# The impact of vitamin D supplementation on mortality rate and clinical outcomes of COVID-19 patients: A systematic review and meta-analysis

**DOI:** 10.1101/2021.01.04.21249219

**Authors:** Leila Nikniaz, Mohammad Amin Akbarzadeh, Hossein Hosseinifard, Mohammad-Salar Hosseini

## Abstract

**Background:** Several studies have suggested the positive impact of vitamin D on patients infected with SARS-CoV-2. This systematic review aims to evaluate the effects of vitamin D supplementation on clinical outcomes and mortality rate of COVID-19 patients.

**Methods:** A comprehensive search was conducted through the databases of PubMed, Scopus, Web of Knowledge, Embase, Ovid, and The Cochrane Library with no limitation in time and language, until December 16, 2020. The results were screened based on their accordance with the subject. Two independent reviewers selected the eligible studies and the outcomes of interest were extracted. Using the Joanna Briggs Institute (JBI) Critical Appraisal Tools for Randomized Controlled Trials (RCTs) and Quasi-Experimental Studies, the remaining results were appraised critically. Statistical analysis was performed using the Comprehensive Meta-Analysis (CMA) software version 2.0.

**Results:** Of the 2311 results, 1305 duplicated results were removed. After screening the titles, abstracts, and the full-text articles of the remaining records, four studies and 259 patients were enrolled, including 139 patients in vitamin D intervention groups. In three of the studies, the patients’ survival and mortality rate were evaluated. The pooled analysis of these studies showed a significantly lower mortality rate among the intervention groups (10.56%) compared with the control groups (23.88%) (OR = 0.264, 95% CI = 0.099–0.708, p-value = 0.008). Two of the studies reported the clinical outcomes based on the World Health Organization’s Ordinal Scale for Clinical Improvement (OSCI) score for COVID-19, where both of them showed a significant decrease in OSCI score in the vitamin D intervention groups. Additionally, One study reported a lower rate of intensive care unit (ICU) admission, and one study reported a significant decrease in serum levels of Fibrinogen.

**Conclusion:** Prescribing vitamin D supplementation to patients with COVID-19 infection seems to decrease the mortality rate, the severity of the disease, and serum levels of the inflammatory markers. Further studies are needed to determine the ideal type, dosage and duration of supplementation.

## Introduction

With the global widespread of the Severe acute respiratory syndrome coronavirus 2 (SARS-CoV-2) and the progressive rise in infection and mortality toll, efficient and effective management of the current medical emergency has become an absolute priority [1]. Great efforts have been taken in most of the involved countries to develop a comprehensive therapeutic approach to prevent and cure the disease. Considering the absence of definite treatment, the high number of the infected people, limited capacity of the healthcare centers, and extensive costs of providing treatments, the researchers and the clinicians are struggling to present appropriate clinical approaches with favorable cost-benefit outcomes, which can help in both preventing the disease and treat the patients, along with lowering the burden of disease on the community [2, 3]. Since no safe medication has been developed yet, the search for the beneficial use of currently-available drugs has been prioritized. Therefore, different ways of improving the immune system’s function have turned to a primary research goal.

One of the potential candidates is vitamin D; a fat-soluble micronutrient that could possibly facilitate the function of the immune system [4, 5]. Although the emerging evidence is growing, a potential association between vitamin D deficiency and severe outcomes in patients with COVID-19 have been reported thus far [6, 7]. The immunomodulatory role of vitamin D has been investigated in the treatment of upper respiratory infections, even before the current pandemic [8]. Vitamin D and related metabolites have a regulatory role in the immune system, thanks to the common receptors found in various innate immune system cells [9, 10]. Furthermore, it can suppress the adaptive immune response in the affected areas (e.g., lung epithelial cells) and, consequently, avoid the pro-inflammatory agents’ harm to prevent further damage [10]. Also, vitamin D plays a protective role against the direct damage of the inflammatory factors – which are secreted during the viral diseases – by lowering the expression rate of inflammatory factors, stimulating the expression of anti-inflammatory factors, and more importantly stimulating the proliferation of the immune cells and their products [11].

Since vitamin D has both stimulating and regulating effects on the immune system, it is reasonable to direct more attention to evaluating the in-hospital and home prescription of this supplement [12]. If the positive impact of vitamin D on COVID-19 patients gets confirmed, it can be used as a cheap and widely available therapeutic aid. Considering the probability of the addition of this supplement in treatment guidelines, it may be beneficial to evaluate the possible effects of vitamin D therapy on infected patients.

According to the numerous evidence available and the lack of a systematic review in this field, this systematic review was conducted to evaluate the impact of prescribing vitamin D supplementation on mortality and clinical outcomes of COVID-19 patients.

## Methods

The current systematic review was conducted according to the recommendations of the Preferred Reporting Items for Systematic Reviews and Meta-Analysis (PRISMA) [13]. The research question of the study was based on PICO and is available in Table 1. Figure 1 presents the PRISMA Flow Diagram of the study. The study was approved by the Ethics Committee of Tabriz University of Medical Sciences, Tabriz, Iran.

**Table 1.** Formulated question of the study based on PICO(S)

| Components of PICO | Defined as |
| --- | --- |
| **Population/Patients** | COVID-19 Patients |
| **Intervention** | Vitamin D Supplementation |
| **Control** | No Vitamin D Supplementation |
| **Outcome** | Mortality Rate, Severity of the Clinical Outcomes |
| **Study Design** | Clinical Trials, Quasi-Experimental, and Interventional Pilot Studies |

**Figure 1.**
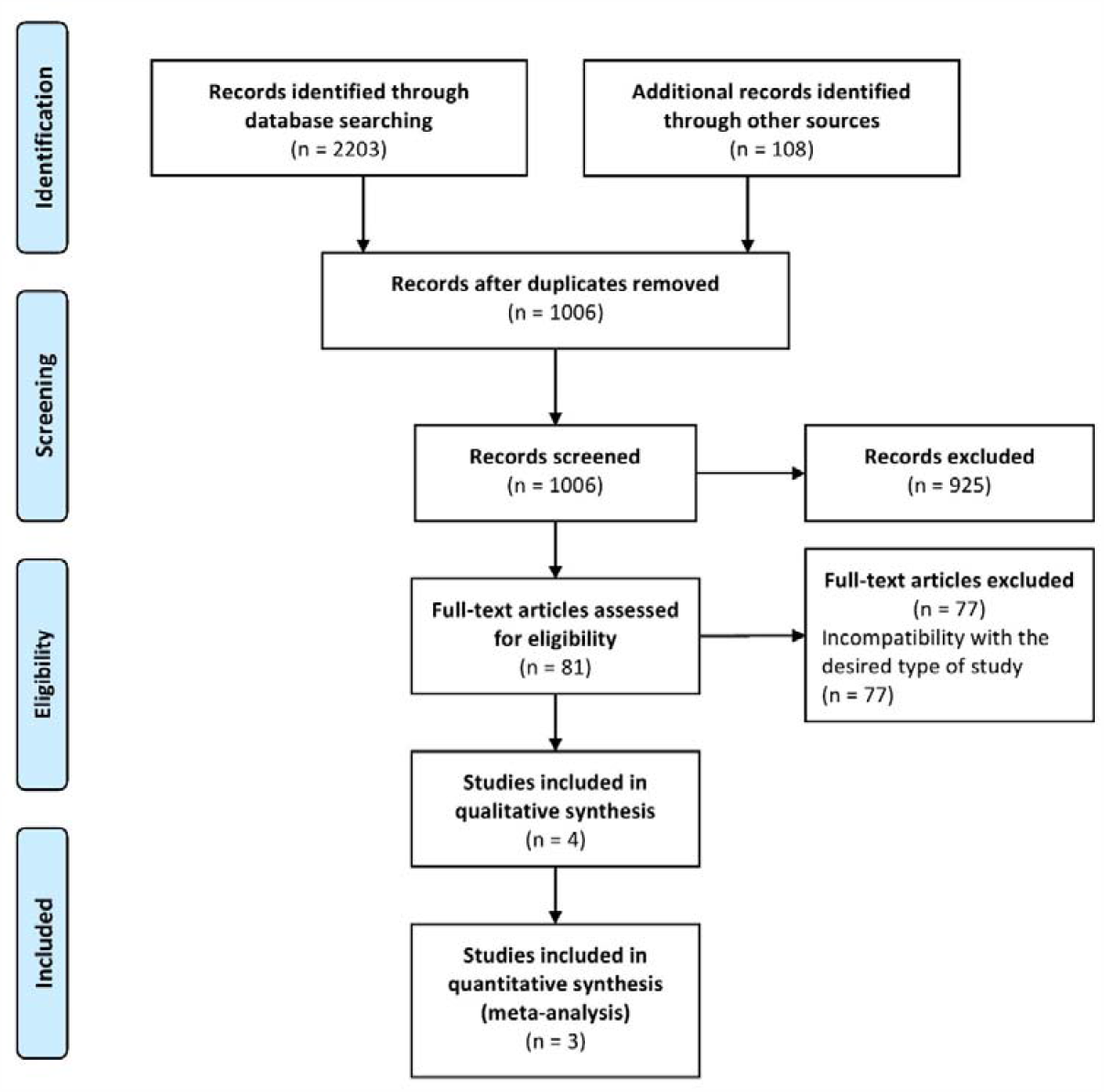
PRISMA Flow Diagram of the study.

### Search Strategy

The current systematic review was designed and conducted in 2020. A comprehensive search was conducted among the databases of PubMed, Scopus, Web of Knowledge, Embase, Ovid, and The Cochrane Library, using a combination of the following free keywords and related MeSH (Medical Subject Headings) Terms: vitamin D, vitamin D3, vit d, cholecalciferol, ergocalciferol, 25-hydroxyvitamin, dihydrotachysterol, calcidiol, 25-hydroxycholecalciferol, covid, covid-19, sars-cov-2, 2019-ncov, coronavirus. The search was conducted on December 16^th^, 2020, without limitation in time and language. Also, in order to increase the accuracy of identifying the related articles, the reference lists of the results were searched, and the related articles were included. The search strategy for PubMed was as follows: (“Vitamin D”[Mesh] OR Vitamin D OR vitamin D3 OR Vit D OR Cholecalciferol OR Ergocalciferol OR 25-Hydroxyvitamin OR Dihydrotachysterol OR calcidiol OR 25-hydroxycholecalciferol) AND ((“COVID-19” [Supplementary Concept]) OR “severe acute respiratory syndrome coronavirus 2” [Supplementary Concept] OR covid OR covid-19 OR sars-cov-2 OR 2019-ncov OR coronavirus).

### Inclusion and exclusion criteria

All clinical trials, quasi-experimental, and pilot studies were included, if they administered any type of vitamin D supplementation to at least one group of confirmed COVID-19-positive patients, regardless of the number of patients, age groups, and language of the studies. Studies without vitamin D intervention and studies other than clinical trials, quasi-experimental, and pilot studies (e.g., case-control studies, cohort studies, observational studies, case reports, reviews, letter to editors, etc.), animal studies, and laboratory studies were excluded.

### Study selection and data extraction

In order to identify the studies relevant to the subject of the review, two researchers independently screened the results by title and abstract, according to their accordance with the study subject and the inclusion/exclusion criteria. Afterwards, the full-text of the remaining records were obtained and assessed for relevancy, and the remaining records were finally included in the study.

Two independent researchers extracted the following data from the included studies: Author(s), year of publication, country of the study, mean ages, characteristics of the populations, type of intervention, and different outcomes.

### Quality assessment

Two independent researchers critically appraised all included studies, using the Joanna Briggs Institute (JBI) Critical Appraisal Tools for Randomized Controlled Trials (RCTs) and Quasi-Experimental Studies.

### Statistical analyses

Statistical analysis was performed on the effect of vitamin D supplementation on the mortality rate, data of which was available in three of the studies, using the Comprehensive Meta-Analysis (CMA) software version 2.0. The odds ratio (OR) was calculated for each of the studies by events and total numbers of patients in two groups. The degree of heterogeneity was defined as significant with a p-value less than 0.05 or I^2^ over 50%. The pooled OR and the 95% confidence interval (95% CI) were calculated, and a p-value less than 0.05 was considered as statistically significant for outcomes. The results were presented as forest plot.

## Results

### Study selection

Of the 2311 results, 1305 were removed due to the duplicity among different sources. After screening the titles and abstracts, 925 more studies were excluded due to incompatibility with the inclusion criteria. After reviewing the full-texts of the remaining records, 77 more records were excluded due to incompatibility of the study design with the inclusion criteria, and finally, four studies were critically appraised for risk of bias and all four were included in the current study. Figure 1 presents the PRISMA Flow Diagram of the study.

### Study characteristics and quality assessment

Two of the studies were appraised by the JBI Critical Appraisal Tools for Randomized Controlled Trials (RCTs) and two of them by the JBI Critical Appraisal Tools for Quasi-Experimental Studies. All the appraised studies are reported as low-risk for bias in terms of having more low-risk domains than high-risk ones. Table 2 and Table 3 present the results of the quality assessment. The results of the risk of bias assessment are present in Table 2 and Table 3.

**Table 2.** Quality assessment of the included quasi-experimental studies.

| Author<br>(year) | Is it clear in the study what is the cause and what is the effect | Were the participants included in any comparisons similar? | Were the participants included in any comparisons receiving similar treatment/care, other than the exposure or intervention of interest? | Was there a control group? | Were there multiple measurements of the outcome both pre and post the intervention/exposure? | Was follow up complete and if not, were differences between groups in terms of their follow up adequately described and analyzed? | Were the outcomes of participants included in any comparisons measured in the same way? | Were outcomes measured in a reliable way? | Was appropriate statistical analysis used? | Overall appraisal |
| --- | --- | --- | --- | --- | --- | --- | --- | --- | --- | --- |
| **C.**<br>**Annweiler**<br>**et al.**<br>**(2020)** | Y | Y | Y | Y | Y | Y | Y | Y | Y | Include |
| **G.**<br>**Annweiler**<br>**et al.**<br>**(2020)** | Y | U | Y | Y | Y | Y | Y | Y | Y | Include |

U: Unclear. N: No. Y: Yes.

**Table 3.** Quality assessment of the included randomized controlled trials.

| Author<br>(year) | Was true randomization used for<br>assignment of participants to treatment<br>groups? | Was allocation to treatment groups<br>concealed? | Were treatment groups similar at the<br>baseline? | Were participants blind to treatment<br>assignment? | Were those delivering treatment blind<br>to treatment assignment? | Were outcomes assessors blind to<br>treatment assignment? | Were treatment groups treated<br>identically other than the intervention of<br>interest? | Was follow up complete and if not, were<br>differences between groups in terms of<br>their follow up adequately described<br>and analyzed? | Were participants analyzed in the<br>groups to which they were randomized? | Were outcomes measured in the same<br>way for treatment groups? | Were outcomes measured in a reliable<br>way? | Was appropriate statistical analysis<br>used? | Overall appraisal |
| --- | --- | --- | --- | --- | --- | --- | --- | --- | --- | --- | --- | --- | --- |
| **Castillo**<br>**et al.**<br>**(2020)** | Y | U | Y | N | U | U | Y | Y | Y | Y | Y | Y | Include |
| **Rastogi**<br>**et al.**<br>**(2020)** | Y | U | Y | Y | U | U | Y | U | Y | Y | Y | Y | Include |

U: Unclear. N: No. Y: Yes.

A total of 259 individuals (140 females and 119 males) were present in the included studies, 139 of which were allocated to the intervention groups. Two of the studies were conducted in France [14, 15], one in India [16] and one in Spain [17]. From the four included studies, Three of the studies reported the mortality rate and survival, two of them reported the clinical outcomes based on the World Health Organization’s Ordinal Scale for Clinical Improvement (OSCI) score for COVID-19 [18], one of them reported the rate of intensive care unit (ICU) admission, and one of the studies reported the changes of the inflammatory markers. Among the included studies, two studies were conducted among a population of aged patients (mean ages 87.7 and 88), and one study among vitamin D deficient patients (25(OH)D < 20 ng/mL). Bolus vitamin D3 was prescribed in three of the studies, two of them along with antibiotics, Hydroxychloroquine, and Corticosteroids (single-dose, oral route, 80.000 IU for one day), and one of them among the vitamin D deficient COVID-19 patients (oral route, 60.000 IU for seven day). In the study with vitamin D deficient patients, oral Cholecalciferol (60,000 IU) was prescribed for 7 days. The supplementation was continued to 14 days for the patients who did not reach the treatment goal of 25(OH)D > 50 ng/mL in the first seven days. From the baseline of 25(OH)D3 = 8.6 [7.1 – 13.1] (ng/ml), the levels of 25(OH)D3 were increased to over 50 in ten patients after seven days of supplementation, and over 50 in two more patients after an overall of 14 days of supplementation. Along with Hydroxychloroquine and Azithromycin, oral Calcifediol was prescribed in one study by the following protocol: 0.532 mg on the day of admission, 0.266 mg on third and seventh days, and then 0.266 mg once every week. The characteristics of the included studies are presented in Table 4.

**Table 4.** Characteristics and information of the included studies.

| Author (Year) | Country | Study population<br>(Total Number:<br>Female/Male) | Age (Mean ±<br>SD) | Number of patients in the<br>Intervention group<br>(Female:Male) | History of<br>diseases –<br>intervention<br>group | History of<br>diseases –<br>control group | Type of<br>intervention | Other administered drugs | Admission<br>to ICU |  | Mortality |  | Results |
| --- | --- | --- | --- | --- | --- | --- | --- | --- | --- | --- | --- | --- | --- |
|  |  |  |  |  |  |  |  |  | Intervention group | Control group | Intervention group | Control group |  |
| C. Annweiler et al.[14]<br>(2020) | France | COVID-19-positive<br>elderly nursing-<br>home residents<br>(66:51/15) | 87.7 ± 9.3 | 57 (45:12) | NA | NA | Vitamin D3<br>(80,000 IU, oral,<br>single dose) | Corticosteroids,<br>Hydroxychloroquine,<br>Antibiotics<br>(Azithromycin/Rovamycin) | NA | NA | 10/57 | 5/9 | The mortality rate was<br>significantly lower in the<br>intervention group (17.5% against<br>55.56 in the control group) (p =<br>0.023). The adjusted HR for<br>mortality according to vitamin D3<br>supplementation was 0.11, and the<br>survival was longer in the<br>intervention group. The OSCI<br>score was lower in the intervention<br>group. |
| M. Castillo et al.[17]<br>(2020) | Spain | COVID-19<br>hospitalized patients<br>(76:31/45) | 53 ± 10 | 50 (23:27) | Hypertension<br>(11/50),<br>cardiac<br>disease (2/50),<br>immunosuppre-<br>ssion/transplan-<br>tation (6/50),<br>lung disease<br>(4/50),<br>diabetes (3/50) | Hypertension<br>(15/26),<br>cardiac<br>disease (1/26),<br>immunosuppre-<br>ssion/transplan-<br>tation (1/26),<br>lung disease<br>(2/26),<br>diabetes (5/26) | Calcifediol (0.532<br>mg, oral, single<br>dose on the day of<br>admission,<br>continued with<br>half dose (0.266<br>mg) on day 3 and<br>7, and then<br>weekly) | Hydroxychloroquine (400 mg<br>every 12 h on the first day, and<br>200 mg every 12 h for the<br>following 5 days),<br>Azithromycin (500 mg orally<br>for 5 days. | 1/50 | 13/26 | 0/50 | 2/26 | The rate of ICU-admission in the<br>intervention group (2%) was lower<br>than the control group (50%) (p <<br>0.001). No death was reported in<br>the intervention group, while two<br>of the control group patients died<br>during their admission in the ICU. |
| **A. Rastogi et al.[16] (2020)** | India | Asymptomatic or mildly symptomatic SARS-CoV-2 RNA-positive patients with vitamin D deficiency (25(OH)D < 20 ng/ml) (40:20/20) | NA (Median 50.0 in the intervention group and 47.5 in the control group) | 16 (10:6) | NA | NA | Cholecalciferol (60,000 IU, oral, daily for 7 days) | Standard care (not specified) | NA | NA | NA | NA | More patients tend to get rid of SARS-CoV-2 after receiving Cholecalciferol ($p < 0.018$) with a significant reduction in fibrinogen level ($p = 0.007$) An overall decrease was observed in the inflammatory markers, but the changes of CRP, Procalcitonin, Ferritin, and D-dimer were not significant |
| **G. Annweiler et al.[15] (2020)** | France | Patients hospitalized for COVID-19 in a geriatric unit (77:38/39) | $88 \pm 5$ | 16 (5:11) | Cancer (4/16)<br>Hypertension (10/16)<br>Cardiac disease (11/16) | Cancer (13/32)<br>Hypertension (21/32)<br>Cardiac disease (18/32) | Vitamin D3 (80,000 IU, oral, single dose, within a few hours of the diagnosis of COVID-19.) | Antibiotics (Quinolones/Beta-lactams/Sulfonamides/Macrolides/Lincosamides/Aminoglycosides), Systemic Corticosteroids, Pharmacological treatments of Respiratory disorders (Beta-2-adrenergic agonists, inhaled corticosteroids, antihistamines, etc.) | NA | NA | 3/16 | 10/32 | Initial bolus vitamin D3 supplementation was associated with less severe clinical outcome and better overall survival rate among the frail elderly. Considering the control group as the reference ($HR = 1$), the fully-adjusted HR for 14-day mortality was 0.37 ($p = 0.28$) for the intervention group. |

COVID-19: Coronavirus disease-2019. CRP: C-reactive protein. HR: Hazard ratio. ICU: Intensive care unit. NA: Not available. OSCI: Ordinal Scale for Clinical Improvement. SARS-CoV-2: Severe acute respiratory
syndrome coronavirus 2.

### Mortality rate and survival

Three of the included studies reported the mortality rate. In the study of C. Annweiler et al. the mortality rate was 55.56% in the control group which was significantly (p = 0.023) higher than the intervention group (17.75%) during the follow-up time (36.6 ± 17 days) [14]. Hazard ratio (HR) for mortality in elderly COVID-19 patients, in accordance with the use of bolus vitamin D3 supplements was HR = 0.11 [95% CI: 0.03-0.48], which indicated that vitamin D supplementation is strongly more effective against mortality, compared with other interventions of the same study, including the use of corticosteroids (HR = 6.64), use of Hydroxychloroquine (HR = 15.07), use of antibiotics (HR = 0.36), and hospitalization (HR = 0.38). In another study, the mortality rate was 7.69% in the control group which was higher than the intervention group (0%), and the patients in the intervention group were all discharged without complications [17]. In the study of G. Annweiler et al., the mortality rate was 31.25% in the control group which was insignificantly (p = 0.28) higher than the intervention group (18.75%), showing a hazard ratio of 0.37 for 14-day mortality [15].

As presented in Figure 2, a meta-analysis was performed on the three studies reporting the mortality rate. The heterogenicity among the studies was not significant (Q = 1.514, df = 2, I^2^ = 0.000, p-value = 0.469). The intervention group consisted of 123, and the control group consisted of 67 patients. Based on the results of the meta-analysis, vitamin D supplementation was associated with a significant reduction in the odds of mortality, compared with the control group (pooled OR =0.264, 95% CI = 0.099-0.708, p-value = 0.008).

**Figure 2.**
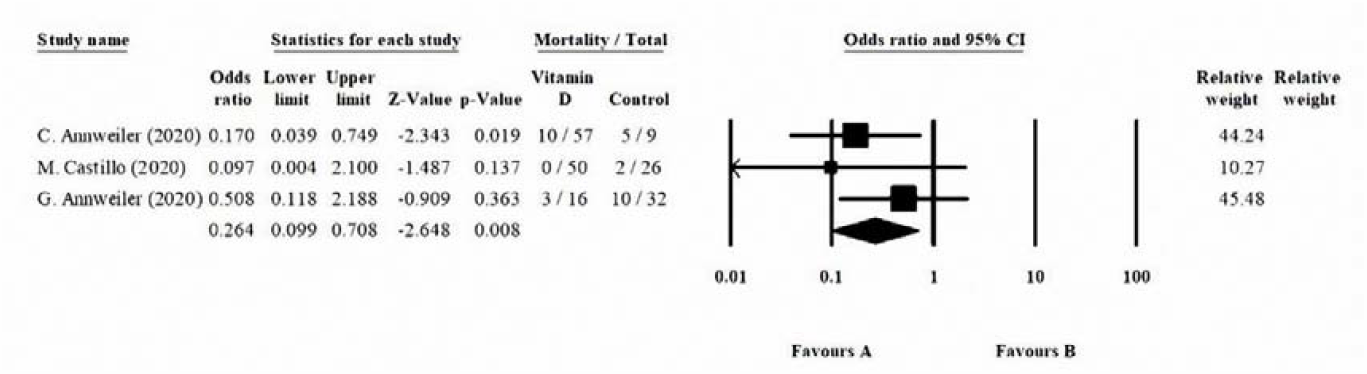
Effect of vitamin D supplementation on the mortality rate of COVID-19 patients.

### Intensive Care Unit (ICU) admission

Only one study reported the Intensive Care Unit (ICU) admission [17]. The Intensive Care Unit (ICU) admission rate was 50% in the control group, in contrast with 2% in the intervention group. In this study, after performing a multivariate logistic regression analysis and adjusting the possible confounding effects on the admission to the ICU, an odds ratio (OR) of 0.03 (95% CI: 0.003 - 0.25) was observed, in favor of no need for ICU admission.

### Secondary outcomes and severity of the disease

The World Health Organization’s Ordinal Scale for Clinical Improvement (OSCI) score for COVID-19 [18] was considered as a secondary outcome in two of the studies [14, 15]. The OSCI score was adjusted for participants’ characteristics, and a significant decrease in the score was observed associated with the bolus supplementation of vitamin D (p = 0.001). The severity of disease was also assessed by the serum levels of inflammatory markers in one of the studies[16]. Unlike C-reactive protein (CRP), Procalcitonin, and D-dimer, the level of Fibrinogen was significantly decreased among the intervention group after the study duration (p-value = 0.007).

## Discussion

To the best of our knowledge, this is the first systematic review exploring the effect of vitamin D supplementation on the mortality rate and clinical outcomes of COVID-19 patients. This review was conducted in order to clarify the effect of vitamin D supplementation on the clinical outcomes of the patients with COVID-19 infection, including the mortality rate, the severity of the disease, and the need for intensive care. Four studies [14-17], including 259 patients with COVID-19 infection, were included and assessed. Our analysis indicated that vitamin D supplementation could positively affect the mortality rate of COVID-19 patients. Moreover, vitamin D supplementation can significantly enhance the patients’ survival, reduce the clinical complications, decrease ICU admission rate, and lower the serum levels of the inflammatory markers.

Vitamin D is a micronutrient which has been investigated vastly due to its unique physiological impacts (i.e., regulating the endocrinological processes, the Renin-Angiotensin-Aldosterone-System (RAAS) pathway, etc.), and regulating the different pathways of the both innate and acquired immune system [19], which would eventually lead to anti-inflammatory, antioxidant, and antiviral characteristics [20, 21]. Considering the immunologic aspects of these characteristics, numerous studies have been conducted in order to investigate the effects of vitamin D on different bacterial and viral infections. Acute upper and lower respiratory infections caused by viral pathogens are among the most researched infections in this regard [22]; and the effects of vitamin D level on the viral diseases including influenza, respiratory syncytial virus, Dengue fever, Hepatitis C and HIV have been investigated in multiple studies for each of these pathogens [23-27]. However, despite the general regulatory role of vitamin D, studies are reporting different results, leading to controversy about the clinical outcomes of vitamin D; for instance, in case of influenza, despite the studies that showed significant results in favor of vitamin D effectiveness in prevention and treatment of the influenza [28, 29], some studies showed no significant benefit in the prescription of vitamin D, like the study carried out in Vietnam, where 1641 children with influenza were randomized to vitamin D and placebo groups. Eventually, no decrease in influenza cases was observed among the group receiving vitamin D [30].

Among the included studies of this review, three studies reported the mortality rate, all showing decrease in mortality rate of the intervention groups. Considering individually, the decrease in mortality rate was significant in only one of the studies. However, the pooled analysis indicated a solid and significant decrease in the mortality rate among patients with vitamin D supplementation. Although the results are completely in favor of vitamin D, decreasing the mortality rate, an ultimate conclusion cannot be drawn due to the lack of studies. Also, the heterogenicity could be affected by the low number of studies. Moreover, some factors in the included studies could affect the patients’ outcomes; two studies were conducted on the aged people, which might affect the mortality rate due to the presence of more comorbidities among the studied population. Similarly, one study was conducted on vitamin D-deficient patients which may show higher effectiveness of intervention, compared with an average population. Another factor that might affect the results is the difference in the dose of supplementation, which is inevitable due to the number of studies.

In contrast to the low number of clinical trials conducted about the efficacy of vitamin D prescription in COVID-19 patients, there is a significant number of observational studies available, which evaluate the association of vitamin D level and the patients’ outcomes, justify to clarify the research path for further evaluation by conducting randomized clinical trials. Most of these studies have indicated a significant association between vitamin D deficiency and severe clinical outcomes of COVID-19 [31, 32]. Age, sun exposure, and diet are the main risk factors for vitamin D deficiency and considering these factors, several studies have been conducted to evaluate the possible link between these factors and the clinical outcomes of COVID-19 [33-35].

Both young and aged people are prone to vitamin D deficiency, which indicates that regardless of the existence of comorbidities, correcting the deficiency could be vital for all patients’ health [36]. However, the difference between these groups is in the etiology of the deficiency, which is due to lack of sun exposure and inadequate dietary intake among the young people, but rather physiologic among the aged people. Hence, both groups are disposed to the risk of severe outcomes of COVID-19. However, it may be challenging to establish the association, solely considering the vitamin D deficiency in the elderly group since a variety of confounding factors are present because of the comorbidities the elderly group may have. Basically, increased age is an independent risk factor for both the vitamin D deficiency and severe outcomes of COVID-19 [37, 38]. The study conducted by lips et al., where the mean levels of vitamin D were measured in the populations of 40 countries and care home residents, mostly consisted of the elderly, have shown a deficiency of over 50% [39]. The sun exposure is another determinant of the vitamin D storage, and with lack of exposure, the deficiency is likely expected. Multiple ecological studies have shown that countries with higher latitude and decreased vitamin D levels have an increased infection rate and poor outcomes [40]. The study conducted by Rhodes et al. demonstrated that countries below 35 degrees north have relatively low mortality and the people above that degree may suffer excess mortality because of the insufficient sunlight to produce adequate vitamin D during winter. This might be indicative of a possible role of vitamin D deficiency in determining poor outcomes [41, 42].

Apart from the etiology, the deficiency has a great impact on the strength of the immune system and therefore, leads to poor outcomes of COVID-19. A retrospective, multi-centric study conducted in the Philippines has demonstrated that in a population of 212 COVID-19 patients, patients with higher vitamin D levels had better outcomes [43]. Vitamin D receptors in the nuclei membrane regulate several defensive proteins and receptors – which are also effective against other viral infections [44]. Receptors recognize the pathogens, like viruses, and the interaction of vitamin D with these receptors could eventually affect the expression of their related genes [45]. In addition to these effects on the innate immune system, it also applies the regulatory impact on the adaptive immune system, which eventually results in inhibition of TH1 proliferation and shifting toward the proliferation of TH2, reduction of the oxidative compounds produced by TH1, influencing the maturation of the T-cells towards the anti-inflammatory subtypes, and prevent the further damage caused by the compounds [19, 46].

Regarding the inflammatory origin of COVID-19 clinical manifestations, it is beneficial further to evaluate inflammatory factors’ role as relative prognostic factors [47]. Previous studies have indicated that several inflammatory markers like C-reactive protein (CRP), Interleukin 6 (IL-6), and erythrocyte sedimentation rate (ESR) have the potential to determine the prognosis and the severity of patients’ clinical outcomes [48-51]. Our study results showed that lower concentrations of these factors have been recorded among the patients who have consumed vitamin D supplementation. In one of the included studies, a significant decrease in Fibrinogen levels was observed after vitamin D supplementation. However, other the decrease in the evaluated inflammatory factors was not statistically significant. This could be explained according to the short follow-up duration and considering the fact that D-dimer, CRP, and ferritin have a relatively longer half-life and might need more time to reflect the impact. Therefore, it is important to determine extended follow-up durations in order to evaluate the paraclinical outcomes better.

The main advantages of the current our systematic review are the rapid synthesis of the information – as the first systematic review on this topic, delivering the importance of further studies by highlighting the current clinical gaps, and providing clear instructions for further studies. The search of databases, study selection, study assessment, and data synthesis were based on defined criteria, and performed by two independent contributors, using the proper methodological tools. However, our review had some limitations. Prominently, the number of publications in the topic is low to draw precise conclusions. Moreover, although the reported heterogenicity was low, the results of the meta-analysis on mortality rate might still be affected by the low number of the included studies. Also, due to the lack of studies reporting the ICU admission and clinical outcomes, performing a quantitative analysis (meta-analysis) on these outcomes was impossible. Also, the difference in the dose of the administered supplementations might have affected the results. We strongly recommend performing further studies, especially clinical trials, on the current topic and among different patients’ groups. Although some population-based studies have already shown the higher prevalence of severe outcomes among the vitamin D-deficient patients, still, we have insufficient evidence-based knowledge about the specific effects of vitamin D supplementation of COVID-19 patients, the impact on the infected patients’ survival, mortality rate and disease progression, the possible side-effects, the proper dosage and route of prescription, the duration of the prescription course, and the potential prophylactic effects; which may be the most beneficial and practical application of vitamin D during this medical state of emergency.

## Conclusion

Vitamin D supplementation seems to decrease the mortality rate, the severity of the disease, and the inflammatory markers’ levels among the COVID-19 infected patients, leading to a better prognosis and increased survival rate. More studies should be conducted to determine the optimum dosage and route of vitamin D supplementation and further investigate the potential prophylactic effects.

## Data Availability

Data used for analyses are available in Table 4.

## Acknowledgement

The research protocol of this study was approved and supported by the Student Research Committee, Tabriz University of Medical Sciences (Grant number: 66884).

